# Toxigenic and non-toxigenic *Vibrio cholerae* serogroups co-circulate across multiple drinking water source types during cholera outbreaks in Zamfara State, northwestern Nigeria

**DOI:** 10.64898/2026.07.09.26357630

**Authors:** Oluchukwu Abba, Naibi Mohammed, Rosemary Okoye, Victor Chijioke Ukwaja, Murtala Saidu, Nurudeen Salisu, Yomi Marie Carole Nyandjou, Umar Abubakar

## Abstract

**Background:** Cholera remains a recurrent public health emergency in Zamfara State, northwestern Nigeria, where communities depend predominantly on untreated and poorly protected water sources. Environmental water bodies serve as reservoirs for *Vibrio cholerae*, sustaining transmission cycles between outbreaks. Despite the severity of recurrent outbreaks in the region, data on the molecular characteristics and serogroup distribution of *V. cholerae* across different drinking water source types in Zamfara State remain critically limited.

**Methodology/Principal Findings:** A cross-sectional environmental surveillance study was conducted between 13 October and 26 November 2025 across five cholera-affected Local Government Areas (LGAs) of Zamfara State: Gusau, Bungudu, Talata Mafara, Zurmi, and Shinkafi. A total of 142 water samples were collected from five source types — rivers, boreholes, wells, tap water, and sachet water. Presumptive isolation was performed on Thiosulfate-Citrate-Bile Salts-Sucrose (TCBS) agar following alkaline peptone water enrichment. Fifty-five presumptive isolates underwent PCR-based molecular confirmation and serotyping using three gene targets: *ompW* (species confirmation, 588 bp), *ctxA* (O1 toxigenicity marker, 302 bp), and *tcpA* (O139 colonisation factor, 120 bp). Presumptive *V. cholerae* was recovered from 55 of 142 samples (38.7%; 95% CI: 30.5–47.3%), with well water recording the highest positivity rate (69.7%; 95% CI: 51.3–83.7%). A statistically significant association was observed between water source type and presumptive *V. cholerae* occurrence (χ² = 23.11, df = 4, p < 0.001). Molecular analysis confirmed 29 isolates (52.7%; 95% CI: 39.2–66.0%) as *V. cholerae*, comprising 22 O1 serotypes (75.9%), one O139 serotype (3.4%), and six non-O1/non-O139 serotypes (20.7%). Toxigenic O1 strains were detected across all five LGAs and in all five water source types, including commercially packaged sachet water. The O139 serotype was identified in a single well-water isolate from Zurmi LGA, representing the first environmental detection of this serotype in Zamfara State.

**Conclusions/Significance:** The co-circulation of toxigenic O1, O139, and non-toxigenic non-O1/non-O139 *V. cholerae* serogroups across five distinct drinking water source types confirms that community water environments serve as genetically diverse reservoirs sustaining cholera transmission in Zamfara State. These findings underscore the urgent need for integrated water quality surveillance, sanitation infrastructure investment, and sustained molecular monitoring of environmental *V. cholerae* populations.

**Author Summary:** Cholera is a severe diarrhoeal disease that kills tens of thousands of people every year, mostly in countries where clean drinking water is difficult to access. It is caused by a bacterium called *Vibrio cholerae*, which can survive and spread through rivers, wells, boreholes, and even commercially packaged drinking water. In Zamfara State, northern Nigeria, cholera outbreaks have returned repeatedly in recent years, yet very little scientific information exists about which types of *V. cholerae* are present in the water people drink every day.

In this study, we collected 142 water samples from five different types of drinking water source — rivers, boreholes, wells, tap water, and sachet water — across five cholera-affected Local Government Areas (LGAs) of Zamfara State. Using a highly sensitive DNA test called PCR, we identified the specific strains of *V. cholerae* present and determined whether they were capable of causing disease.

We found disease-causing *V. cholerae* in all five types of water source, including sachet water that people commonly assume is safe. We also made the first-ever detection of a particularly dangerous strain type, called O139, in Zamfara State. Our findings show that no single water source in these communities can be regarded as reliably free of contamination, and that regular water quality monitoring, sanitation improvements, and targeted public health interventions are urgently needed to protect communities from cholera across Zamfara State and similar settings in sub-Saharan Africa.

## Introduction

Cholera is an acute secretory diarrhoeal disease caused by the Gram-negative bacterium *Vibrio cholerae*, transmitted primarily through the ingestion of contaminated water or food. The disease remains a major global public health challenge, particularly in low- and middle-income countries where access to safe drinking water, adequate sanitation, and proper hygiene is limited [1,2]. Cholera outbreaks are strongly associated with inadequate water infrastructure and are frequently exacerbated by environmental factors such as flooding, which facilitate contamination of water sources and rapid inter-community spread [3]. In Nigeria, cholera continues to pose a recurrent public health threat, particularly in northern regions where access to potable water and functional sanitation infrastructure remains chronically deficient [4,5].

*Vibrio cholerae* is autochthonous to aquatic environments including rivers, lakes, estuaries, and groundwater systems where it persists as part of the natural microbial community [6,7]. These environmental reservoirs sustain inter-epidemic persistence of the pathogen and represent a critical interface between environmental contamination and human exposure across diverse water source types. Transmission occurs predominantly via the faecal–oral route, underscoring the primacy of water quality surveillance in cholera epidemiology [8]. In endemic settings such as Nigeria, contaminated surface water, shallow wells, boreholes, and even packaged water have been identified as sources of infection, establishing the imperative for multi-source environmental monitoring [9,10].

Based on the lipopolysaccharide (O) antigen, *V. cholerae* is classified into over 200 serogroups; however, only O1 and O139 cause epidemic and pandemic cholera [11,12]. The O1 serogroup comprising Classical and El Tor biotypes is responsible for the ongoing seventh pandemic, while O139 emerged in the early 1990s as the first non-O1 strain capable of epidemic spread [13]. Non-O1/non-O139 *V. cholerae* (NOVC) strains are widely distributed in environmental waters and are increasingly recognised as emerging human pathogens capable of causing gastroenteritis, wound infections, and septicaemia [14,15]. Critically, some NOVC strains carry the toxin-coregulated pilus gene (*tcpA*) a key colonisation factor and the receptor for the CTXφ bacteriophage that carries the cholera toxin genes conferring the potential to evolve into toxigenic strains through horizontal gene transfer [13,16].

Zamfara State has experienced recurrent cholera outbreaks in recent years, with communities relying on a diverse range of water sources including untreated surface water, hand-dug wells, boreholes, piped tap water, and commercially packaged sachet water for domestic use. The simultaneous use of multiple source types each with distinct vulnerability profiles to environmental contamination creates a complex epidemiological landscape in which *V. cholerae* may co-circulate across different water environments. Despite this public health burden, published data on the environmental occurrence, virulence gene profiles, and serogroup distribution of *V. cholerae* across different drinking water source types in Zamfara State remain scarce, constraining risk stratification, outbreak response, and the design of targeted interventions.

This study therefore aimed to investigate the presence, molecular characterisation, and serogroup distribution of *Vibrio cholerae* in five types of drinking water source across five cholera-affected LGAs in Zamfara State, Nigeria, using PCR-based detection of *ompW* (species identification), *ctxA* (O1 toxigenicity marker), and *tcpA* (O139 colonisation factor). Characterising the co-circulation and environmental distribution of both toxigenic and non-toxigenic serogroups across diverse water source types is prerequisite to evidence-based cholera surveillance and water safety intervention in this endemic setting.

## Materials and Methods

### Ethical approval

Ethical clearance was obtained from the Zamfara State Ministry of Health Research Ethics Committee prior to sample collection and laboratory analysis. Community and local government authority consent was obtained at each sampling site. No human subjects, patient data, or biological specimens from human participants were involved.

### Study area

This cross-sectional environmental surveillance study was conducted across five LGAs of Zamfara State, northwestern Nigeria: Gusau (state capital), Bungudu, Talata Mafara, Zurmi, and Shinkafi. All five LGAs had documented active cholera outbreaks at the time of sampling. Zamfara State is characterised by a predominantly rural population relying on rivers, hand-dug wells, boreholes, tap water distribution systems, and commercially packaged sachet water each carrying distinct risks of microbial contamination.

### Sample collection

The study was conducted between 13 October and 26 November 2025. Purposive sampling was adopted to target locations at elevated cholera transmission risk within each LGA. Water samples were collected from five source types: rivers, boreholes, wells, tap water, and sachet water. All samples except sachet water were collected aseptically in pre-labelled 100 mL sterile bottles. Sachet water samples were purchased in original sealed packaging. A total of 142 water samples were transported to the Microbiology Laboratory, Federal University Gusau, within 2–4 hours of collection for immediate processing. Variation in sachet water brand availability across LGAs resulted in a shortfall of eight samples from the planned total; this reflects real-world water-access conditions rather than a methodological constraint.

### Enrichment and isolation

One millilitre of each water sample was inoculated into 9 mL of sterilised alkaline peptone water (APW) and incubated at 37 °C for 18 hours. Enriched cultures were plated on Thiosulfate-Citrate-Bile Salts-Sucrose (TCBS) agar and incubated at 37 °C for 24 hours. Yellow (sucrose-fermenting) colonies characteristic of *V. cholerae* were recorded as presumptive positives and subcultured on fresh TCBS agar for 24 hours at 37 °C to obtain pure cultures.

### DNA extraction

Fifty-five presumptive isolates were inoculated into Cary-Blair transport medium and transported to Bioformatics Services molecular research laboratory, Ibadan, for confirmatory PCR-based analysis. Genomic DNA was extracted using the ZR Fungal/Bacterial DNA Miniprep kit (Zymo Research, USA) following the manufacturer’s protocol. Briefly, 2 mL of bacterial cell broth was homogenised in a ZR BashingBead™ Lysis Tube at maximum speed for 5 minutes. The lysate was centrifuged at >10,000 × g for 1 minute; up to 400 µL of supernatant was processed through sequential spin columns with pre-wash and wash buffers, and DNA was eluted in 100 µL of DNA Elution Buffer. DNA concentration and purity were assessed by NanoDrop spectrophotometry prior to PCR amplification.

### PCR amplification and gene targets

PCR was performed in 25 µL reactions containing 12.5 µL Taq 2× Master Mix (NEB, M0270), 1 µL each of 10 µM forward and reverse primer, 2 µL DNA template, and 8.5 µL nuclease-free water. Three gene targets were amplified: (i) *ompW* for species confirmation (588 bp): F-5’CACCAAGAAGGTGACTTTATTGTG-3’; R-5’GAACTTATAACCACCCGCG-3’, the *ompW* outer membrane protein gene is highly conserved across *V. cholerae* serogroups and rarely amplified in other bacteria, making it the most reliable species-level confirmation target [17]; (ii) *ctxA* for O1 toxigenicity (302 bp): F-5’CTCAGACGGGATTTGTTAGGCACG-3’; R-5’TCTATCTCTGTAGCCCCTATTACG-3’ encodes the A subunit of cholera toxin; (iii) *tcpA* for O139 classification (120 bp): F-5’AGAAGAACACGATAAGAAAACCG-3’; R-5’CGAATCAATCGCACGCTG-3’ encodes the major structural subunit of the toxin-coregulated pilus (TCP), an essential colonisation factor and receptor for the CTXφ bacteriophage [13,19]. Thermocycling: initial denaturation 94 °C for 5 min; 36 cycles of 94 °C for 30 s, 64 °C for 30 s, 72 °C for 45 s; final extension 72 °C for 7 min; hold 10 °C. Amplicons were resolved on 2% agarose gels and visualised under UV transillumination [18].

### Molecular identification and serotyping

Among *ompW*-confirmed *V. cholerae* isolates: (a) *ctxA*-positive isolates were classified as O1 serotype; (b) *tcpA*-positive/*ctxA*-negative isolates were classified as O139 serotype; and (c) isolates negative for both *ctxA* and *tcpA* were classified as non-O1/non-O139. This classification follows established PCR-based serotyping protocols [17,20].

### Drinking water quality standards

Results were interpreted with reference to the World Health Organization (WHO) Guidelines for Drinking-water Quality [1] and the Nigerian Standard for Drinking Water Quality (NSDWQ, NIS 554:2015), both of which specify zero tolerance for *V. cholerae* in water intended for human consumption.

### Data analysis

Data were analysed using descriptive statistics (frequencies, percentages, and proportions with 95% Wilson score confidence intervals). A chi-square (χ²) test of independence was applied to assess the association between water source type and occurrence of presumptive *V. cholerae* isolates. Fisher’s Exact Test (Freeman-Halton extension for r×c tables) was used for secondary analyses where expected cell counts fell below five. Statistical significance was set at p < 0.05. All proportions are reported with 95% Wilson score CIs.

## Results

### Distribution of water samples

A total of 142 water samples were collected from five LGAs and five water source types (Table 1). Borehole water constituted the largest proportion of samples (42/142; 29.6%), followed by well water (33/142; 23.2%), river water (27/142; 19.0%), sachet water (22/142; 15.5%), and tap water (18/142; 12.7%). Gusau LGA contributed the most samples (n = 30; 21.1%); Talata Mafara contributed the fewest (n = 26; 18.3%) owing to limited sachet water brand availability in cholera-affected communities within that LGA.

**Table 1.** Distribution of water samples across five LGAs and five source types, Zamfara State, Nigeria (n = 142)

| LGA | River | Borehole | Tap | Well | Sachet | Total n | % |
| --- | --- | --- | --- | --- | --- | --- | --- |
| Gusau | 6 | 6 | 6 | 6 | 6 | 30 | 21.1 |
| Bungudu | 8 | 8 | 0 | 8 | 5 | 29 | 20.4 |
| Zurmi | 6 | 6 | 6 | 6 | 5 | 29 | 20.4 |
| Shinkafi | 3 | 10 | 6 | 5 | 4 | 28 | 19.7 |
| Talata Mafara | 4 | 12 | 0 | 8 | 2 | 26 | 18.3 |
| **Total n** | **27** | **42** | **18** | **33** | **22** | **142** | **100** |

Tap water was absent from Bungudu and Talata Mafara LGAs within the sampled communities. Sachet water availability was limited in Talata Mafara (n=2 brands) and Shinkafi (n=4 brands). LGA = Local Government Area.

### Presumptive Vibrio cholerae isolates

Of the 142 samples analysed, 55 (38.7%; 95% CI: 30.5–47.3%) yielded presumptive *V. cholerae* on TCBS agar (Table 2). Well water recorded the highest presumptive positivity rate (23/33; 69.7%; 95% CI: 51.3–83.7%), followed by river water (13/27; 48.1%; 95% CI: 29.4–67.5%), tap water (5/18; 27.8%; 95% CI: 11.1–52.5%), borehole water (10/42; 23.8%; 95% CI: 12.6–39.5%), and sachet water (4/22; 18.2%; 95% CI: 6.1–39.2%). The association between water source type and presumptive *V. cholerae* occurrence was statistically significant (χ² = 23.11, df = 4, p < 0.001), confirming that contamination rates differed significantly across the five source types (Table 3).

**Table 2.** Presumptive *Vibrio cholerae* isolates by LGA and water source type with positivity rates and 95% confidence intervals.

| LGA | River | Borehole | Tap | Well | Sachet | Total +ve | Sampled | % |
| --- | --- | --- | --- | --- | --- | --- | --- | --- |
| Gusau | 4 | 0 | 0 | 3 | 1 | 8 | 30 | 26.7 |
| Bungudu | 1 | 3 | 0 | 5 | 0 | 9 | 29 | 31.0 |
| Zurmi | 4 | 3 | 3 | 4 | 1 | 15 | 29 | 51.7 |
| Shinkafi | 1 | 2 | 2 | 3 | 1 | 9 | 28 | 32.1 |
| Talata Mafara | 3 | 2 | 0 | 8 | 1 | 14 | 26 | 53.8 |
| **Total +ve** | **13** | **10** | **5** | **23** | **4** | **55** | **142** | — |
| **Positivity (%)** | 48.1 | 23.8 | 27.8 | 69.7 | 18.2 | 38.7 | — | — |
| **95% CI** | 29.4–67.5 | 12.6–39.5 | 11.1–52.5 | 51.3–83.7 | 6.1–39.2 | 30.5–47.3 | — | — |

95% CIs computed using the Wilson score method. +ve = presumptive positive (yellow colony on TCBS agar).

**Table 3.** Chi-square test of independence: water source type and occurrence of presumptive *Vibrio cholerae* (n = 142)

| Water source | Positive n (%) | Negative n (%) | Total n | $\chi^2$ | df | p |
| --- | --- | --- | --- | --- | --- | --- |
| River | 13 (48.1) | 14 (51.9) | 27 |  |  |  |
| Borehole | 10 (23.8) | 32 (76.2) | 42 |  |  |  |
| Tap water | 5 (27.8) | 13 (72.2) | 18 |  |  |  |
| Well | 23 (69.7) | 10 (30.3) | 33 |  |  |  |
| Sachet | 4 (18.2) | 18 (81.8) | 22 |  |  |  |
| **Total** | **55 (38.7)** | **87 (61.3)** | **142** | **23.11** | **4** | **<0.001** |

Significance threshold: $p < 0.05$ .

### Molecular confirmation and serotyping

Of the 55 presumptive isolates, 29 (52.7%; 95% CI: 39.2–66.0%) were confirmed as *V. cholerae* by amplification of the species-specific *ompW* gene at 588 bp (Fig 1). Serotyping revealed that toxigenic and non-toxigenic serogroups co-circulate across multiple water source types and LGAs (Table 4): 22 isolates were O1 serotype (*ctxA*-positive; 75.9%; 95% CI: 57.2–88.5%); one isolate was O139 serotype (*tcpA*-positive; 3.4%; 95% CI: 0.2–17.8%); and six isolates were non-O1/non-O139 (20.7%; 95% CI: 8.9–39.7%). Gel images confirming amplification of *ctxA* (302 bp) and *tcpA* (120 bp) are presented in Fig 2 and Fig 3, respectively.

**Fig 1.**
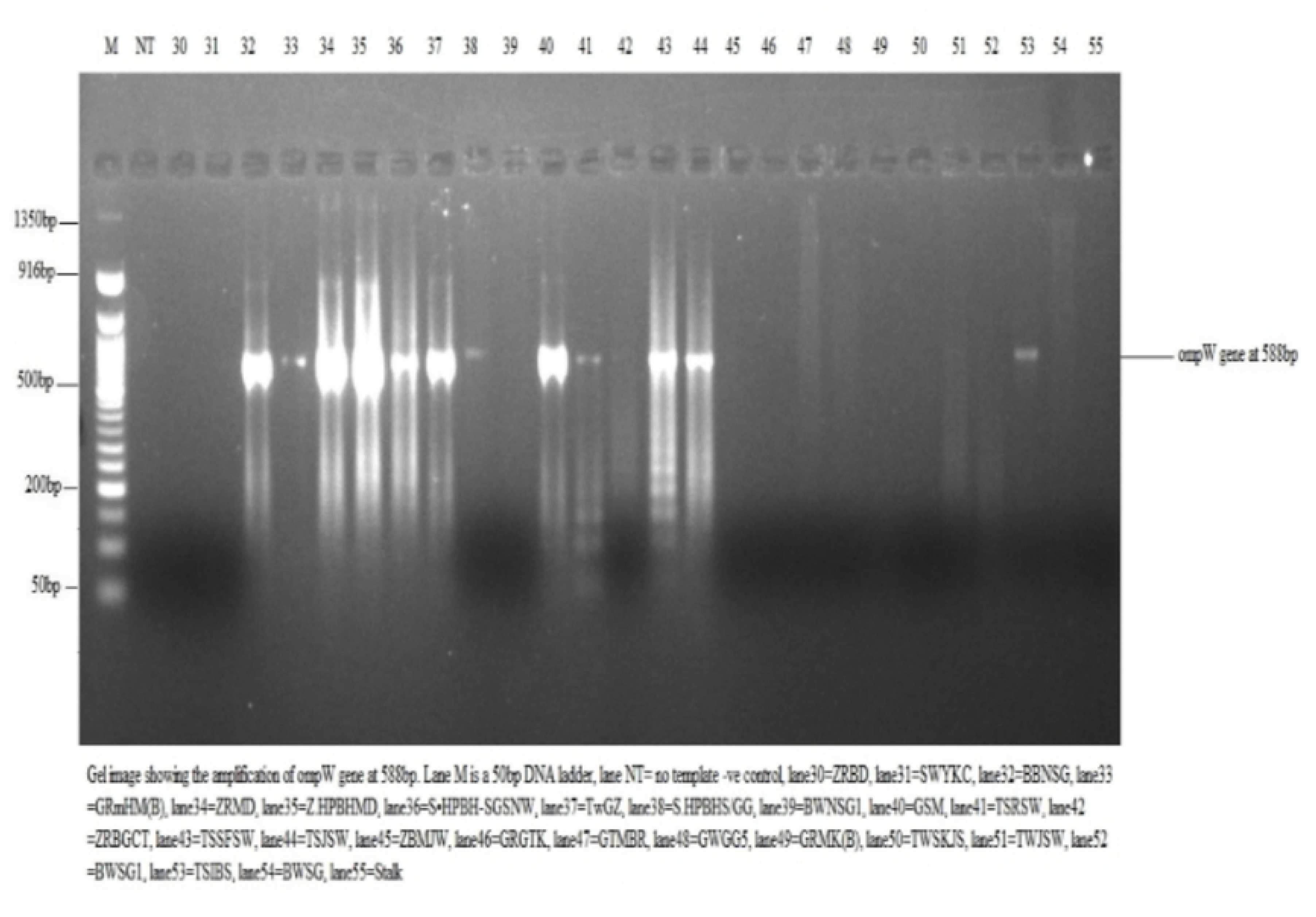
Agarose gel electrophoresis showing amplification of *ompW* gene at 588 bp.

**Fig 2.**
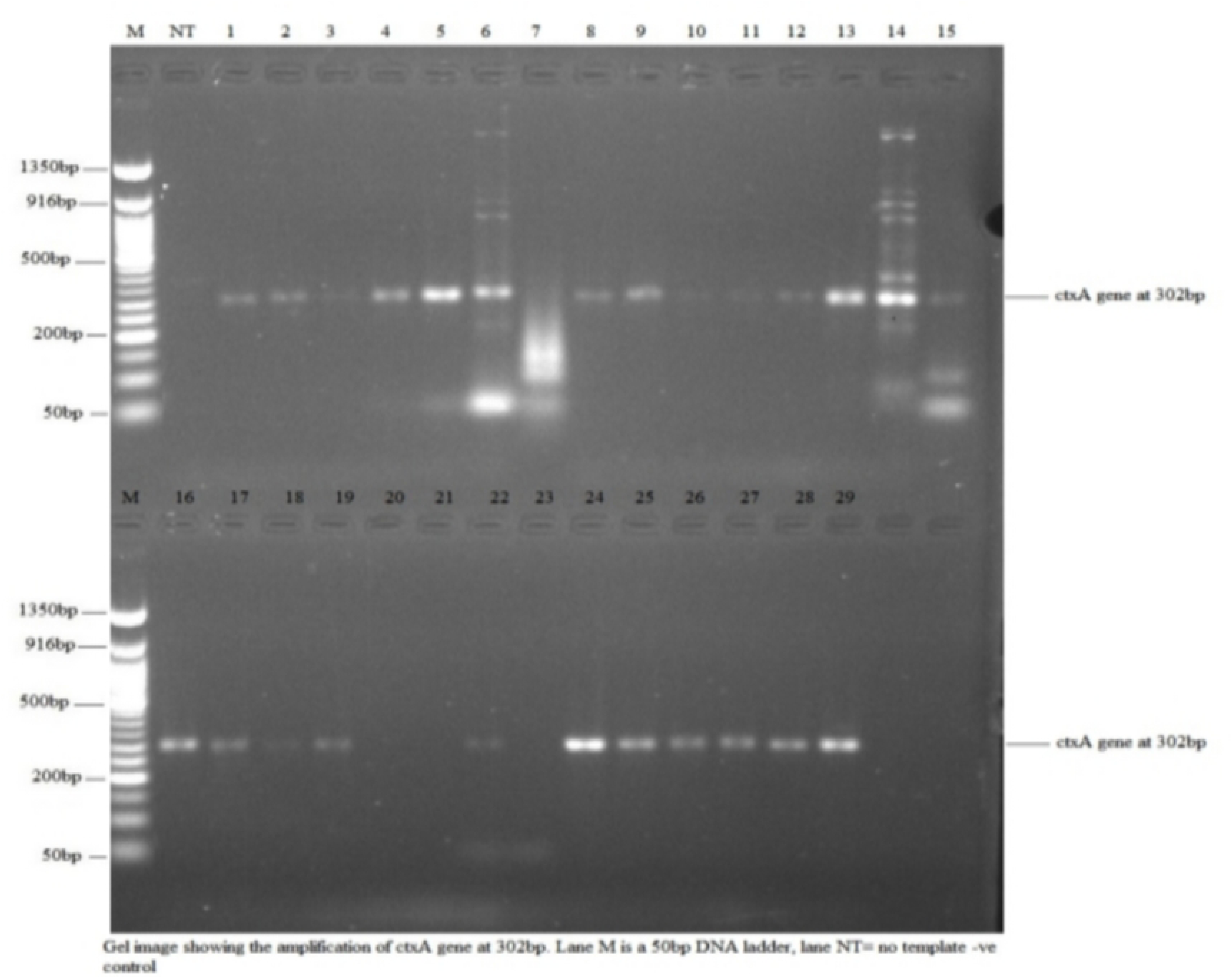
Agarose gel electrophoresis showing amplification of *ctxA* gene at 302 bp.

**Fig 3.**
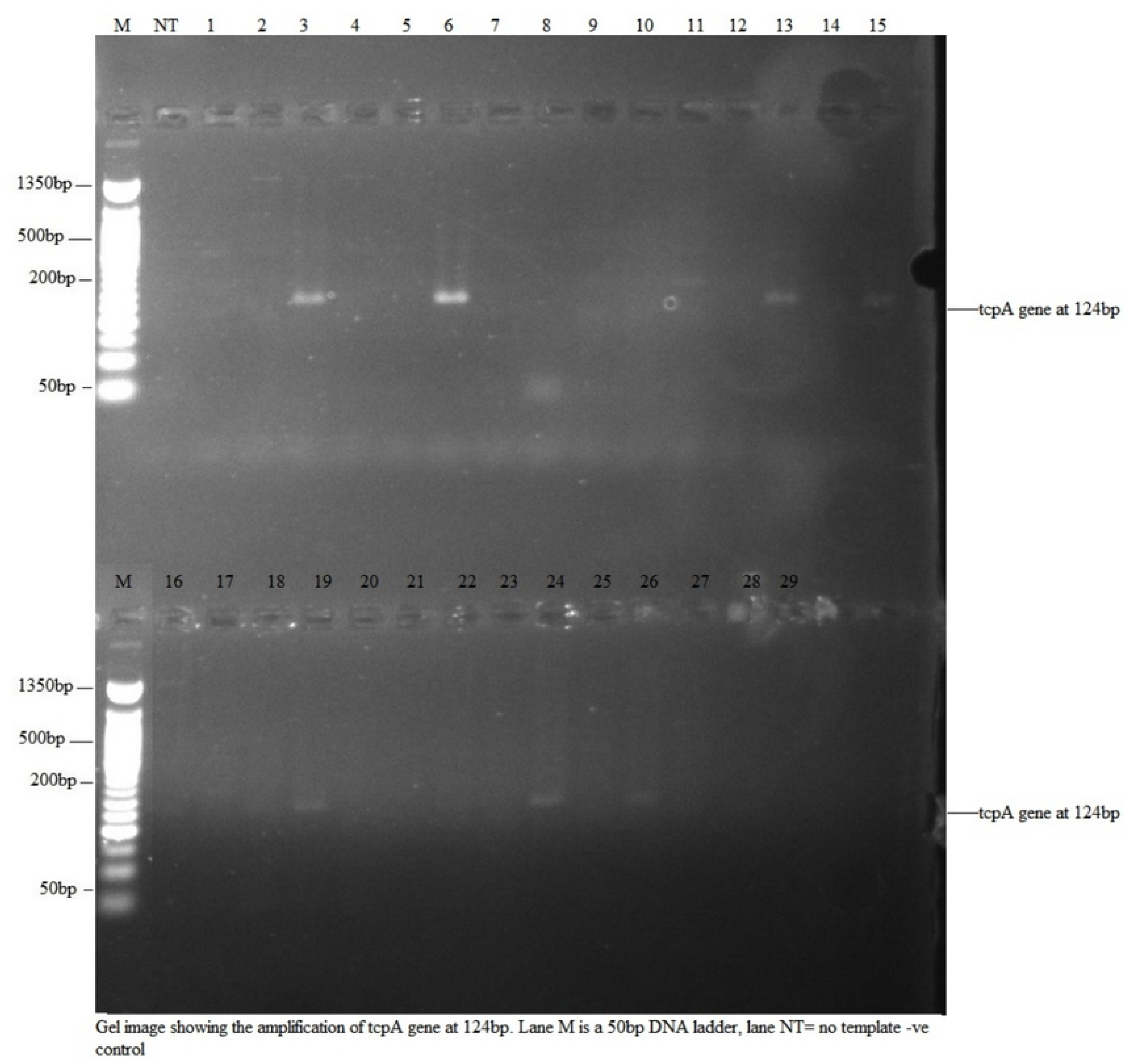
Agarose gel electrophoresis showing amplification of *tcpA* gene at 120 bp.

**Table 4.** PCR-based molecular confirmation and serotyping results for *Vibrio cholerae* isolates (n = 55 tested; n = 29 confirmed)

| Gene target | Amplicon (bp) | Purpose | n/N positive (%) | 95% CI | Serotype classification |
| --- | --- | --- | --- | --- | --- |
| *ompW* | 588 | Species confirmation | 29/55 (52.7%) | 39.2–66.0% | *V. cholerae* confirmed |
| *ctxA* | 302 | Cholera toxin — O1 marker | 22/29 (75.9%) | 57.2–88.5% | O1 (toxigenic) |
| *tcpA* | 120 | TCP — O139 marker | 1/29 (3.4%) | 0.2–17.8% | O139 (toxigenic) |
| Neither *ctxA* nor *tcpA* | — | Non-toxigenic classification | 6/29 (20.7%) | 8.9–39.7% | Non-O1/non-O139 |

Denominators: *ompW* percentage is of 55 tested; remaining percentages are of 29 *ompW*-confirmed *V. cholerae*. 95%
CIs: Wilson score method. TCP = toxin-coregulated pilus.

### Distribution of O1 and O139 serogroups

The O1 serotype was detected across all five LGAs and in all five water source types, demonstrating the broadest environmental distribution (Table 5). Talata Mafara LGA yielded the highest number of O1 isolates (n = 10); all confirmed isolates from this LGA were toxigenic (100%). The O139 serotype was identified in a single well-water isolate from Zurmi LGA (ZWBABT), representing the first documented environmental detection of this serotype in Zamfara State.

**Table 5.** Distribution of confirmed *Vibrio cholerae* O1 and O139 serogroups by LGA and water source type.

| LGA | River | Borehole | Tap | Well | Sachet | O1 total | O139 total |
| --- | --- | --- | --- | --- | --- | --- | --- |
| Gusau | 0 | 0 | 0 | 0 | 1 (GSM) | 1 | 0 |
| Bungudu | 0 | 1 (BBSG2) | 0 | 1 (BWNSG2) | 0 | 2 | 0 |
| Zurmi | 0 | 1 (ZHPBHMD) | 1 (ZTKK) | 3 (ZWSDG, ZWCSDG, ZWZBT) | 0 | 5 | 1 (ZWBABT) |
| Shinkafi | 1 (SSKTR) | 0 | 1 (STMDW) | 1 (SWGLD) | 1 (SSHG) | 4 | 0 |
| Talata Mafara | 3 (TSRSW, TSSFSW, TSJSW) | 1 (TSMBHS) | 0 | 5 (TWGGS, TWGH, TWGCL, TWKGS, TWGZ) | 1 (TSIBS) | 10 | 0 |
| **Total** | **4** | **3** | **2** | **10** | **3** | **22** | **1** |

O1 defined by *ctxA* positivity (302 bp). O139 defined by *tcpA* positivity (120 bp), *ctxA*-negative. Sample codes in Supplementary Table S1.

### Distribution of non-O1/non-O139 serogroups

Non-O1/non-O139 *V. cholerae* isolates were detected in Bungudu, Zurmi, and Shinkafi LGAs, recovered from river, borehole, and sachet water (Table 6). Zurmi showed the widest diversity, with non-O1/non-O139 isolates spanning three source types. Although lacking *ctxA* and *tcpA*, environmental non-O1/non-O139 populations frequently carry alternative virulence determinants — including RTX toxin, haemolysin A (*hlyA*), ToxR transcriptional regulator, and Type VI Secretion System components — associated with pathogenesis and potential to acquire cholera toxin genes through horizontal gene transfer [21,22,23].

**Table 6.** Distribution of non-O1/non-O139 *Vibrio cholerae* serogroups by LGA and water source type.

| LGA | River | Borehole | Sachet | Total non-O1/non-O139 |
| --- | --- | --- | --- | --- |
| Bungudu | 1 (BRN1) | 1 (BBNSG) | 0 | 2 |
| Zurmi | 1 (ZRMD) | 1 (ZBKK) | 1 (ZSBSK) | 3 |
| Shinkafi | 0 | 1 (S.HPBH.SGSNW) | 0 | 1 |
| **Total** | **2** | **3** | **1** | **6** |

Classification: *ompW*-positive, *ctxA*-negative, *tcpA*-negative.

### Virulence profile by LGA

Overall, 23 of 29 confirmed isolates (79.3%) were toxigenic. Talata Mafara and Gusau LGAs yielded exclusively toxigenic isolates; Zurmi showed the greatest serotypic diversity, with O1, O139, and non-O1/non-O139 strains co-detected across river, borehole, tap water, and well sources within the same LGA (Table 7). Freeman-Halton Exact Test indicated that the proportion of toxigenic isolates did not differ significantly across the five LGAs (p = 0.211), a finding attributable to the limited statistical power afforded by small per-LGA confirmed isolate counts (range: n = 1–10). Similarly, serotype distribution across water source types was not statistically significant (3×5 table: χ² = 8.40, df = 8, p = 0.396; toxigenic vs. non-toxigenic 2×5 table: χ² = 7.16, df = 4, p = 0.128).

**Table 7.** Virulence profile of molecularly confirmed *Vibrio cholerae* isolates by LGA: toxigenic versus non-toxigenic classification.

| LGA | O1 (ctxA+) n | O139 (tcpA+) n | Non-O1/non-O139 n | Total n | % Toxigenic | Fisher's p* |
| --- | --- | --- | --- | --- | --- | --- |
| Gusau | 1 | 0 | 0 | 1 | 100.0 | 1.000 |
| Bungudu | 2 | 0 | 2 | 4 | 50.0 | 0.180 |
| Zurmi | 5 | 1 | 3 | 9 | 66.7 | 0.339 |
| Shinkafi | 4 | 0 | 1 | 5 | 80.0 | 1.000 |
| Talata Mafara | 10 | 0 | 0 | 10 | 100.0 | 0.068 |
| **Total** | **22** | **1** | **6** | **29** | **79.3%** | **p = 0.211 (NS)** |

Toxigenic = O1 + O139 combined. % Toxigenic = $(O1 + O139) / \text{Total confirmed} \times 100$ . \*Pairwise Fisher's Exact $p$ = each LGA tested against all remaining LGAs combined, two-sided. Overall $p$ = Freeman-Halton Exact Test for the $5 \times 2$ contingency matrix. NS = not statistically significant ( $p > 0.05$ ). Non-significance reflects limited statistical power at per-LGA sample sizes (range: $n = 1-10$ ).

## Discussion

This study provides the first PCR-confirmed, multi-source environmental surveillance of *Vibrio cholerae* serogroups across five cholera-affected LGAs in Zamfara State, Nigeria. The co-circulation of toxigenic O1, the epidemically significant O139, and non-toxigenic non-O1/non-O139 serogroups simultaneously across five distinct drinking water source types directly informs both risk communication and the design of targeted water safety interventions.

### Differential contamination across water source types

The statistically significant association between water source type and *V. cholerae* occurrence (χ² = 23.11, df = 4, p < 0.001) confirms that contamination risk is strongly structured by source type. Well water exhibited the highest presumptive positivity rate (69.7%; 95% CI: 51.3–83.7%), reflecting the inherent vulnerability of shallow, poorly protected groundwater to faecal contamination through pit latrine seepage, surface runoff, and inadequate well-head protection. Hydrogeological investigation in neighbouring Sokoto State confirmed coliform contamination of shallow aquifers linked to pit latrines and septic systems [24], consistent with the Kampala outbreak investigation in which unprotected well water contaminated by faecal runoff was the primary transmission vehicle [25]. River water was the second most contaminated source type (48.1%; 95% CI: 29.4–67.5%), consistent with the established role of surface water as a recipient of diffuse faecal contamination [26,27].

Crucially, *V. cholerae* was detected in tap water from Zurmi and Shinkafi LGAs despite these sources theoretically benefiting from centralised treatment, and in commercially packaged sachet water across multiple LGAs. The detection of toxigenic O1 in sachet water, a source consumers frequently regard as safe, is particularly concerning and may reflect production-level contamination or post-production handling failures. The detection of *V. cholerae* across all five water source types collectively demonstrates that no single source category within these communities is reliably free from contamination risk, consistent with the recent global systematic review by Awere-Duodu *et al.* [10] confirming substantial prevalence of *V. cholerae* in both surface water and groundwater systems worldwide.

### Co-circulation of three serogroups

The simultaneous detection of O1, O139, and non-O1/non-O139 serogroups in Zurmi LGA, their co-detection across multiple water source types within the same LGA demonstrates the genetic diversity of the *V. cholerae* population circulating in Zamfara State’s water environments. The predominance of O1 (75.9% of confirmed isolates) across all five LGAs aligns with the current global dominance of the El Tor O1 biotype in the seventh pandemic [30]. Aquatic environments serve as evolutionary crucibles where gene exchange between strains is facilitated by the proximity and persistence of diverse bacterial populations [35], creating conditions conducive to the emergence of novel toxigenic lineages through horizontal gene transfer [13,16].

### The O139 detection: novel and epidemiologically significant

The identification of a single O139 isolate (ZWBABT) from well water in Zurmi LGA constitutes the first reported environmental detection of this serotype in Zamfara State. The O139 serogroup demonstrated epidemic potential since its emergence in South Asia in the early 1990s [12,13], and its detection in a West African environmental water source is consistent with genomic surveillance data showing continued O139 circulation outside Asia [28]. Because this isolate carries *tcpA*, the receptor for the CTXφ bacteriophage, it retains the genetic architecture necessary for potential acquisition of cholera toxin genes through horizontal gene transfer, a process that could convert this strain into a fully toxigenic variant [19,16]. Continued molecular monitoring of O139 strains in environmental water sources is therefore warranted.

### Non-O1/non-O139 strains: beyond classical virulence markers

The six non-O1/non-O139 isolates recovered across three LGAs and three water source types including sachet water are not epidemiologically inconsequential. Environmental non-O1/non-O139 *V. cholerae* populations frequently carry alternative virulence determinants including RTX toxin, haemolysin A (*hlyA*), the ToxR transcriptional regulator, mannose-sensitive haemagglutinin pilus (*mshA*), and components of the Type VI Secretion System (T6SS) [21,23]. Westerström *et al.* [22] demonstrated that non-O1/non-O139 strains can exhibit virulence characteristics comparable to clinical isolates despite lacking classical cholera toxin genes. The recovery of these strains from borehole and sachet water sources suggests that the distribution of potentially pathogenic non-toxigenic *V. cholerae* in Zamfara State extends beyond surface water into source types with broader consumer reliance.

### Hotspot LGAs and spatial clustering

Talata Mafara and Zurmi LGAs emerged as epidemiological hotspots. Talata Mafara yielded ten confirmed O1 isolates from four water source types all toxigenic despite having the lowest total sample count, indicating a high environmental burden of toxigenic *V. cholerae* independent of sampling volume. Zurmi was the only LGA where all three serogroups co-circulated simultaneously across multiple source types. Although the Freeman-Halton Exact Test did not detect a statistically significant difference in toxigenic proportions across LGAs (p = 0.211), this reflects limited statistical power at per-LGA confirmed isolate counts (range: n = 1–10) rather than genuine homogeneity. The clustering of presumptive positive isolates within specific wards notably Sabon Gida and Nahuche in Bungudu, and Birnin Tsaba in Zurmi is consistent with localised point-source contamination where multiple households depend on common water bodies, facilitated by surface runoff transporting faecal contaminants into community water sources [26,34,5].

### Public health implications

The co-circulation of toxigenic and non-toxigenic *V. cholerae* serogroups across five drinking water source types — including sources consumers regard as treated or safe — represents a substantial multi-layered public health threat. WHO and NSDWQ standards specify zero tolerance for *V. cholerae* in drinking water; the widespread non-compliance documented here constitutes a public health emergency. Evidence confirms that contamination also occurs after collection during storage in uncovered containers [32,33], compounding the exposure risk beyond source water quality. Targeting well water — the highest-risk source type — through well-head protection, regular disinfection, and community hygiene education should be prioritised as the most immediately impactful intervention, alongside regulatory enforcement of packaged water quality standards.

## Conclusion

This study demonstrated that toxigenic *V. cholerae* O1 — alongside the epidemically significant O139 serotype and non-toxigenic non-O1/non-O139 strains carrying alternative virulence determinants — co-circulate across multiple drinking water source types in five cholera-affected LGAs of Zamfara State, Nigeria. The statistically significant association between water source type and *V. cholerae* contamination (χ² = 23.11, df = 4, p < 0.001), and the disproportionately high contamination of well water (69.7%; 95% CI: 51.3–83.7%), confirm shallow groundwater as the highest-risk exposure pathway. The detection of toxigenic O1 strains in all five water source types and the first environmental detection of O139 in Zamfara State underscore that no single source category is reliably free from contamination. Future studies should incorporate whole-genome sequencing to resolve strain phylogenetics, temporal monitoring to characterise seasonal variation in contamination, and spatial correlation of environmental isolates with clinical outbreak data.

These findings establish an urgent evidence base for integrated water quality surveillance, targeted sanitation investment, and sustained public health interventions across Zamfara State.

## Data Availability

All primary data are provided in the manuscript and Supporting Information. Raw data are available from the corresponding author on reasonable request.

## Supporting Information

**S1 Table. Master list of 55 presumptive isolates with sample codes, LGA, water source, collection location, and molecular result.**

## Declarations

### Ethics statement

Ethical clearance was obtained from the Zamfara State Ministry of Health Research Ethics Committee. Community consent was obtained at each sampling site. No human subjects, patient data, or biological specimens from human participants were involved.

### Author contributions

Conceptualisation: OA; Methodology: OA, NM, RO, VCU, YMCN, UA; Investigation: OA, NM, RO, VCU, MS, NS; Formal Analysis: OA, NM, VCU, MS; Writing – Original Draft: OA; Writing – Review & Editing: all authors; Supervision: OA.

## Acknowledgements

The authors thank Solidarities International for sponsoring the research, Bioformatics Services, Ibadan, for molecular analysis support; the Zamfara State Ministry of Health for ethical clearance, RUWASSA for field access facilitation; and all community and LGA authorities who granted sampling permissions.

